# Ex-vivo mucolytic and anti-inflammatory activity of BromAc in tracheal aspirates from COVID-19

**DOI:** 10.1101/2021.12.23.21268347

**Authors:** Jordana Grazziela A. Coelho dos Reis, Geovane Marques Ferreira, Alice Aparecida Lourenço, Ágata Lopes Ribeiro, Camila Pacheco da Silveira Martins da Mata, Patrícia de Melo Oliveira, Daisymara Priscila de Almeida Marques, Linziane Lopes Ferreira, Felipe Alves Clarindo, Murillo Ferreira da Silva, Heitor Portella Póvoas Filho, Nilson Roberto Ribeiro Oliveira Junior, Maisah Meyhr D’Carmo Sodré, Sandra Rocha Gadelha, George Rego Albuquerque, Bianca Mendes Maciel, Ana Paula Melo Mariano, Mylene de Melo Silva, Renato Fontana, Lauro Juliano Marin, Renata Santiago Alberto Carlos, Amanda Teixeira Sampaio Lopes, Fabrício Barbosa Ferreira, Uener Ribeiro dos Santos, Íris Terezinha Santos de Santana, Hllytchaikra Ferraz Fehlberg, Rachel Passos Rezende, João Carlos T Dias, Eduardo Gross, Gisele Assis Castro Goulart, Marie Gabriele Santiago, Ana Paula Motta Lavigne de Lemos, Aline O da Conceição, Carla Cristina Romano, Luciana Debortoli de Carvalho, Olindo Assis Martins Filho, Claudio Almeida Quadros, Sarah J Valle, David L Morris

**Author notes:** **Corresponding authors and addresses:** Jordana G. Coelho dos Reis. Department of Microbiology, Instituto de Ciências Biológicas, Universidade Federal de Minas Gerais, Av. Antônio Carlos, 6627, Campus Pampulha, CEP: 31270-901, Belo Horizonte, MG, Brazil. and David L. Morris, Mucpharm Pty Ltd Sydney NSW Australia.

## Abstract

COVID-19 is a lethal disease caused by the pandemic SARS-CoV-2, which continues to be a public health threat. COVID-19 is principally a respiratory disease and is often associated with sputum retention, for which there are limited therapeutic options. In this regard, we evaluated the use of BromAc^®^, a combination of Bromelain and Acetylcysteine (NAC). Both drugs present mucolytic effect and have been studied to treat COVID-19. Therefore, we sought to examine the mucolytic, antiviral, and anti-inflammatory effect of BromAc^®^ in tracheal aspirate samples from critically ill COVID-19 patients requiring mechanical ventilation.

**Method:** Tracheal aspirate samples from COVID-19 patients were collected following next of kin consent and mucolysis, rheometry and cytokine storm analysis was performed.

**Results:** BromAc^®^ displayed a robust mucolytic effect in a dose dependent manner. BromAc^®^ showed anti-inflammatory activity, reducing the action of cytokine storm, chemokines including MIP-1alpha, CXCL8, MIP-1b, MCP-1 and IP-10, and regulatory cytokines IL-5, IL-10, IL-13 IL-1RA and total reduction for IL-9 compared to NAC alone and control. BromAc^®^ acted on IL-6, demonstrating a reduction in G-CSF and VEGF-D at concentrations of 125 and 250µg.

**Conclusion:** These results indicate robust mucolytic and anti-inflammatory effect of BromAc^®^ in tracheal aspirates from critically ill COVID-19 patients, indicating its potential as a therapeutic strategy to COVID-19.

## Introduction

On March 11, 2020, the World Health Organization (WHO) declared a pandemic for SARS-CoV-2, a new coronavirus that causes COVID-19 (WHO, 2020). While most persons infected with SARS-CoV-2 remain asymptomatic or oligosymptomatic, a significant proportion of patients may develop the severe form of the disease, which is characterized by systemic multiorgan failure with the pulmonary system being the first and most targeted site. Spike protein (S) from SARS-CoV-2 is recognized by the angiotensin-converting enzyme 2 (ACE 2) receptor, present mainly in respiratory epithelial cells. After viral replication inside cells, the virus is released via exocytosis and spreads quickly and swiftly in the bronchoalveolar epithelium, attracting several immune cells to the infection site and promoting a strong proinflammatory environment and hypersecretion of mucus. SARS-CoV-2 infection induces mucin overexpression further promoting disease. As mucins are critical components of the innate immunity, unravelling their expression profiles that dictate the course of disease could greatly enhance our understanding and management of COVID-19 [1]. In fact, a comprehensive snapshot of blood mucin seems to discriminate symptomatic COVID-19 from patients without disease based on expression of MUC1, MUC2, MUC4, MUC6, MUC13, MUC16 and MUC20 [1].

The S protein, the virion’s binding tool to host cells, becomes an ideal therapeutic target. However, several attempts failed to produce safe and efficient specific direct-acting antiviral therapy. More importantly, studies focusing on drugs that are administered into the airway to act more rapidly at the targeted site are still scarce. In this regard, BromAc^®^ is comprised of a combination of Bromelain and Acetylcysteine, two proteolytic drugs that have been studied as a repurposed agent to treat COVID-19 [2-4]. Moreover, BromAc^®^ is currently used for the treatment of the rare highly mucinous tumor pseudomyxoma peritonei. This combination acts as a biochemical agent capable of destroying glycoprotein S, which makes BromAc^®^ an attractive strategy for promoting local antiviral activity. *In vitro*, it synergistically inhibited the infectivity of two strains of SARS-CoV-2 cultivated in Vero, BGM and CALU-3 cells, showing an antiviral effect of 4 log reduction [4].

Therefore, BromAc^®^ combines the ability of breaking the peptide bonds of the amino acid backbone of mucin, together with disruption of disulphide bonds (SS) between the sugar side chains, causing dissolution of mucins. These mucins are a significant component of the viscous sputum seen in many patients with COVID-19 [5]. Cytokines and chemokines are small peptides which may be glycosylated, making them an attractive target for BromAc^®^. In the present study we sought to examine the mucolytic, antiviral, and anti-inflammatory effect of BromAc^®^ in tracheal aspirate samples from COVID-19 patients. This is the first study of BromAc^®^ in COVID-19 sputum.

## Material and Methods

### BromAc^®^

Lyophilized sterile Bromelain (Mucpharm Pty Ltd) was resuspended in phosphate-buffered saline and aliquots were stored at -20°C. Acetylcysteine (NAC) injection at 200mg/mL was kept ambient until use.

### Patient Samples

Tracheal aspirate (TA) samples (2-10mL) were collected during the early morning routine from 20 COVID-19 patients, aging from 18-80 years-old (male n=12) under mechanical ventilation at the intensive care unit of Hospital de Ilhéus. All patients included in the study tested positive for SARS-CoV-2 by RT-PCR targeting E gene. Only secretion productive patients were included in the study. Samples were aspirated into sterile tracheal secretion collectors and immediately processed in a biosecurity level 3 laboratory. Tracheal aspirate samples were treated with the combination of Bromelain at different concentrations and NAC 2% (20mg/mL) (BromAc^®^) and incubated at 37°C. After incubation, flow through assays, rheometric measurements and cytokine storm assessment were performed. Participant’s next of kin were consented and signed the written informed consent on their behalf to participate in the investigation. This study followed the principles of Helsinki declaration as well as the resolution #466/2012 from the Brazilian Ministry of Health for research involving humans. This investigation was approved by the Santa Cruz State University (UESC) Institutional Brazilian Ethics Committee (Approval number: 45919121.6.0000.5526).

### Flow Through assay

For assessing the impact of BromAc^®^ and its mucolytic effect in the viscosity of tracheal aspirate (TA) samples from COVID-19 patients (n=10), TA specimens were treated with 125 to 250µg of Bromelain and 2% NAC (BromAc^®^) using a mucosal atomization device (MAD nasal) (Teleflex) and incubated at 37°C in 6 well plates. After incubation and visual assessment of TA specimens, time of mucolysis was assessed. Then, treated TA samples were passed through cell strainers (70µm) placed into 50mL falcon polypropylene tubes. Samples were pipetted into cell strainers and volume flow through was collected and measured. Mincing and repeated pipetting was strictly avoided to measure flow through accurately. Volume flow through was harvested, aliquoted and stored at -80°C.

### Rheological measurements

The rheological profile of samples was performed on a DV-III Digital Rheometer (Brookfield, MA, USA), equipped with a CP52 spindle (Cone angle: 3.0°). Three sample groups were evaluated: control (no treatment, n= 10), NAC (treated with n-acetylcysteine, n= 4), BromAc^®^ 125 (treated with Bromelain at 125 µg and NAC 2%, n= 3) and BromAc^®^ 250 (treated with Bromelain at 250 µg and NAC 2%, n= 10). After preparation and addition of the active principles, as performed previously, the samples were homogenized for 1 minute and placed in a water bath at 37 °C for 30 minutes. Subsequently, 0.5 mL of each sample was placed in the sampling cup and subjected to analysis. The equipment was controlled using Rheocalc V3.3 software (Brookfield, MA, USA) programmed to vary the spindle speed from 0.01 to 250 RPM, with an evaluation of 30 points between these values and equilibrium time 30 seconds at each new speed.

### Assessment of Immunological inflammatory mediators in tracheal aspirate samples from COVID-19 patients by Luminex

In order to verify the effect of BromAc^®^ in the inflammatory mediators present in tracheal aspirate (TA) samples from COVID-19 patients, TA specimens were treated with 125 to 250 µg of Bromelain and 2% NAC (BromAc^®^) using the MAD device as described above. Treated and untreated TA specimens were incubated at 37 °C in 6 well plates for 1 hour. After that, TA specimen aliquots were initially cleared by centrifugation at 800 x g for 10 min, at room temperature and TA supernatants were transferred to fresh 2mL microtubes. Samples were diluted 1:10 and incubated with magnetic beads covered with monoclonal antibodies specific to several immunological mediators, such as: chemokines (CXCL8, CCL11, CCL3, CCL4, CCL2, and CXCL10), inflammatory cytokines (IL-1β, IL-6, TNF-α, IL-12, IFN-γ, IL-15 and IL-17), regulatory cytokines (IL-1Ra, IL-9, IL-10) and growth factors (FGF-basic, PDGF, VEGF, G-CSF, GM-CSF, IL-2 and IL-7). Experiments were carried out according to the manufacturer’s instructions using the Procarta Human Cytokine 27-plex Assay, (Invitrogen, CA, USA). The immunological mediators were measured in TA samples and the concentrations of each sample was determined according to standard curves run for each molecule tested using a fifth parameter logistic fit analysis. The results were expressed as pg/mL for all mediators tested.

### Statistical Analysis

All data that followed parametric distribution were analyzed by ANOVA one way followed by post-hoc Dunnet’s multiple comparisons test amongst groups. For correlation analysis, Pearson’s correlation test was employed for data with parametric distribution. For non-parametric distribution data, the Kruskal-Wallis test was applied, followed by the Dunn’s multiple comparisons test. For comparative analysis between two independent groups, the non-parametric Mann-Witney test was used. Spearman correlation test was employed for data with non-parametric distribution. All the analysis were carried out using GraphPad Prism, version 8.0, (San Diego, CA, USA). Significant statistical differences were considered if p value was less or equal 0.05.

## Results

### Determination of mucolytic effect of BromAc^®^ and the impact of cellularity in the in vitro treatment of tracheal aspirate samples from COVID-19 patients

In order to determine whether BromAc^®^ treatment would exert a mucolytic effect on tracheal aspirate samples in a dose-dependent manner, samples were treated with 2% NAC as well as BromAc^®^ 125 and 250 µg. Tracheal aspirate samples were treated for 30 minutes, as described in material and methods. Figure 1A illustrates the mean flow through of samples treated with BromAc^®^ 125 and 250 µg or 2% NAC. Higher mean flow through was observed for samples treated with BromAc^®^, in a dose-dependent manner, providing evidences of its robust mucolytic effect. Flow through recovery was 68% for samples treated with BromAc^®^ 125 µg and over 80% for samples treated with BromAc^®^ 250 µg. The mucolytic effect can also be easily observed in the visual aspect of the samples before and after the treatment with BromAc^®^ 250 µg (Figure 1B). Since the adhesive interactions between cells and other mucus constituents may significantly affect the viscoelasticity of mucus, cellularity analysis (total and live cell count, Figures 1 C-D) of tracheal aspirate samples was assessed after treatment with BromAc^®^ 250 µg. The data demonstrates that cellularity correlates inversely with the mucolytic effect of BromAc^®^ (p= 0.0002, Pearson correlation R^2^ = -0.9880, Figure 1C). In addition, this correlation was seen also with the live cell compartment assessed in all samples BromAc^®^ (p= 0.0009, Pearson correlation R^2^ = -0.9520, Figure 1D). These results likely indicate that the mucolytic effect of BromAc^®^ is seen in samples composed in most of the mucus rather than samples with cell aggregates. New formulations combining cell dissociation components such as DNase or heparin to BromAc^®^ should be considered for future studies.

**Figure 1.**
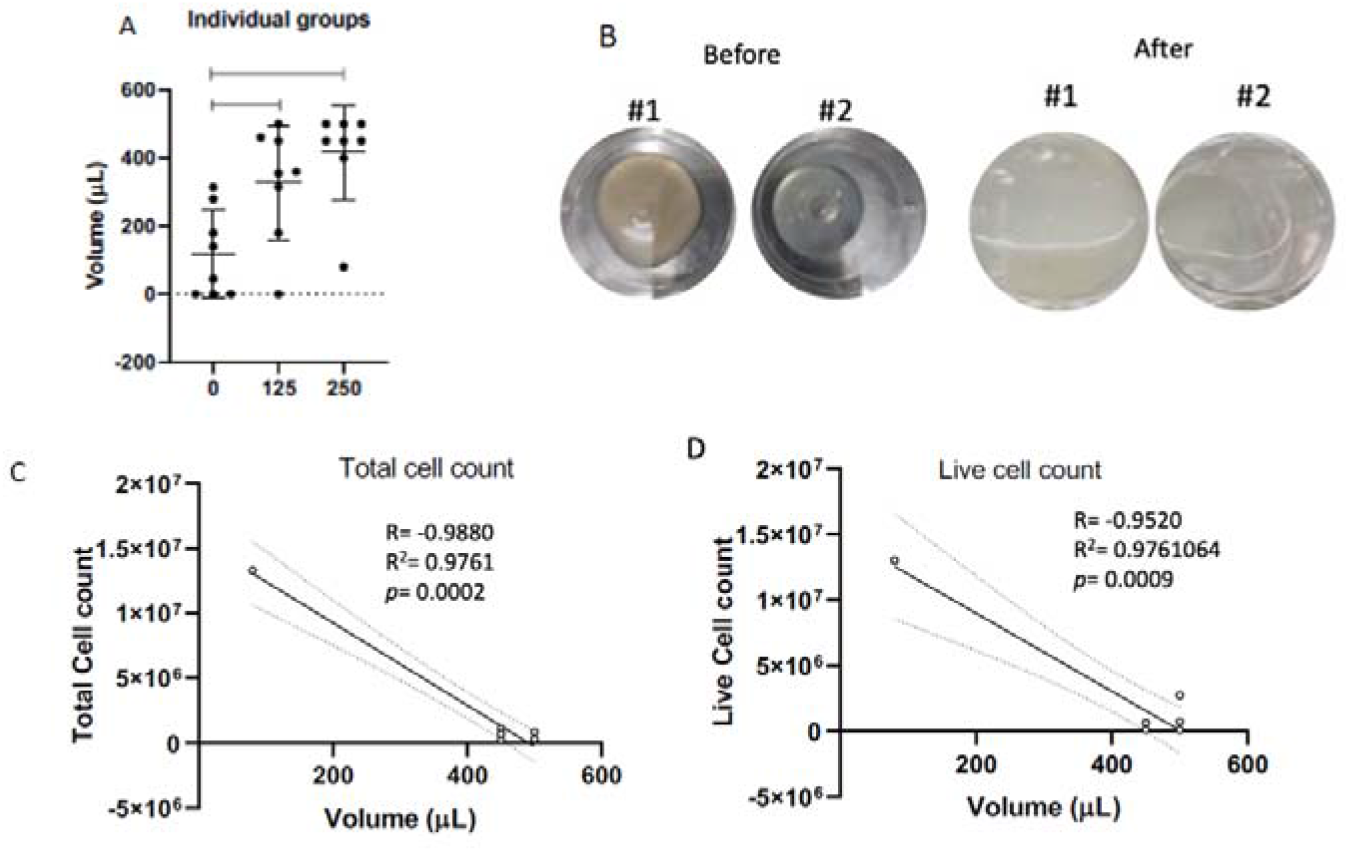
Effect of BromAc^®^ in tracheal aspirate samples from eight COVID-19 patients using flow through method. Volume of flow through in microliters is represented in scatter plots with median and interquartile range for a curve with 0, 125 and 250 µg of Bromelain in addition to 2% NAC (A). Visual aspect of tracheal aspirate sample before and after treatment BromAc^®^ 250μg (B). Correlation of mucolytic effect of BromAc^®^ with tracheal aspirate sample cellularity. XY dispersion graphs demonstrate the results for cellularity including total cell count (C) and live cell count (D) according to volume of flow through in microliters. Tables on the right display the results of Pearson r coefficient, 95% confidence Interval as well as P value and P value summary.

As another method of measuring the impact of BromAc^®^ treatment on the viscosity of tracheal aspirates from COVID-19 patients, we performed rheological measures on samples upon BromAc^®^ treatment. Figure 2 shows the impact of the treatment of tracheal aspirate samples on the viscosity using a rheometer. As can be seen in Figure 2A, the viscosity of tracheal aspirate samples treated with BromAc^®^ 125 or 250 µg reduced significantly from the samples non-treated or treated with 2% NAC. Regarding the rheological profile (Figure 2B), the three samples showed characteristics of a non-Newtonian viscoelastic fluid. Therefore, under low shear, the tracheal aspirate sample behaves like an elastic solid (deformation) and under high shear like a viscous liquid (flow). This rheological signature is characteristic of cross-linked mucin polymers in a mucus, indicating a lightly entangled network [6-8]. However, for samples treated, a greater reduction in viscosity was observed for samples treated with BromAc^®^ 250 µg as clearly demonstrated by scatter plots and heatmap analysis (Figure 2B-D). Heatmap analysis further demonstrated that deformation and mucolysis are accelerated by BromAc^®^ 250 µg (Figure 2D).

**Figure 2.**
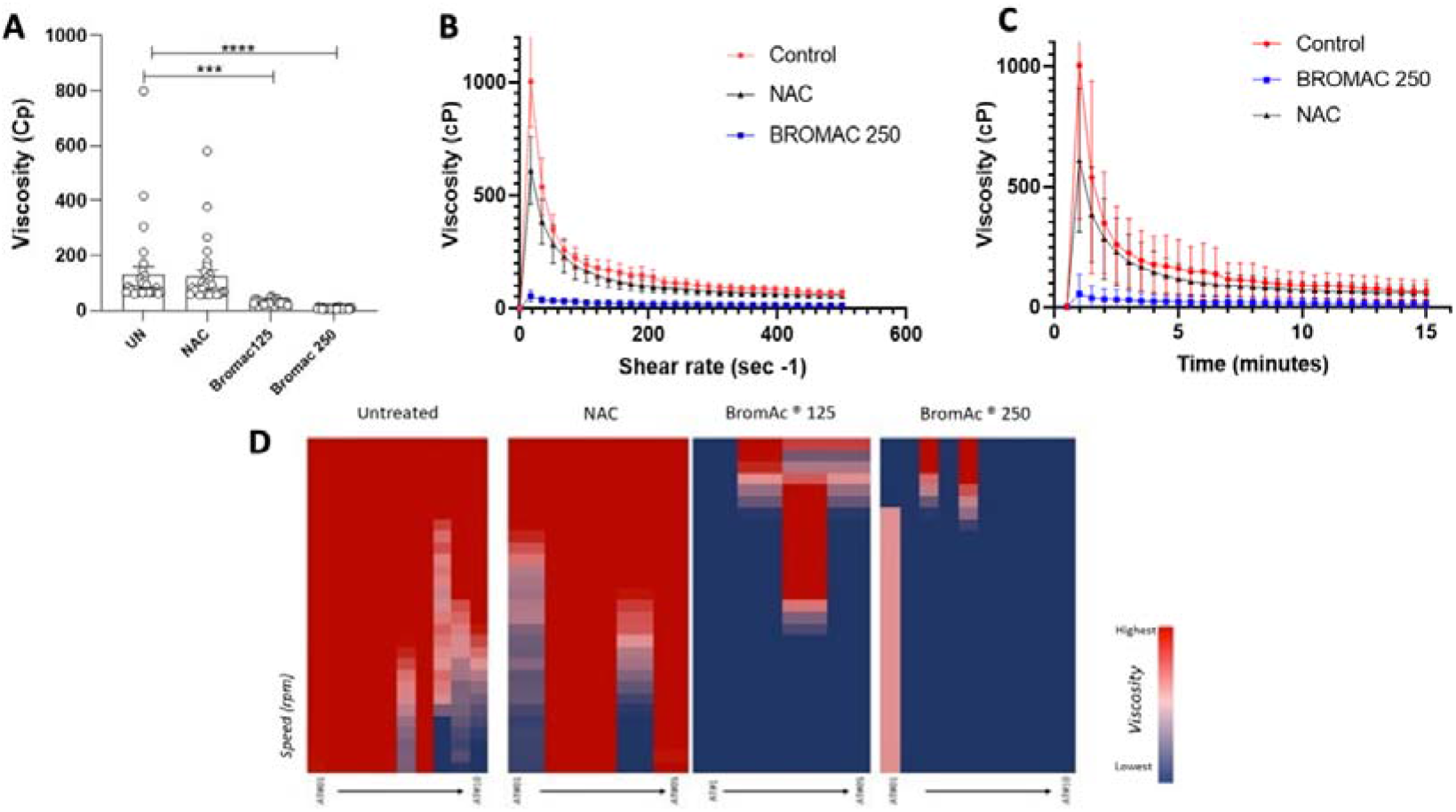
Rheological measurements of tracheal aspirate samples from COVID-19 patients untreated (UN) and after treatment with N-Acetylcysteine (NAC), BromAc^®^ 250μg. Results plotted as scatter graphs over floating bars expressing minimum and maximum as well as the average line (A). Effect on viscosity as function of shear rate expressed in sec-1 (B) and along time expressed in minutes (C). Data are expressed as mean ± SEM. A value of p < 0.05 was considered to indicate statistically significant difference. Heatmap analysis demonstrating viscosity intensity in individual samples. Color key is provided in the figure showing highest (red) and lowest (blue) viscometry (cP) and Speed (rpm) (D). Asterisks identify statistical differences at p values: * p<0.05; ** p<0.01; *** p<0.001; **** p<0,0001.

### Anti-inflammatory activity of BromAc^®^ in tracheal aspirate samples from critically ill COVID-19 patients

In order to further expand BromAc^®^’s effect on the immune components present in the tracheal aspirate samples from COVID-19 patients, the detection of chemokines, cytokines and growth factors was performed after treatment with BromAc^®^. This experiment was performed in similar conditions as the previous curve experiment in which samples were treated with 2% NAC alone as well as BromAc^®^ 125 and 250µg. Tracheal aspirate samples were treated for 60 minutes under the same conditions as the first experiment.

Results show that BromAc^®^ treatment generally and massively abrogated the chemokines (Figure 3). Of note, BromAc^®^ decreased the expression of chemokines CCL2, CCL3, CCL4 and CXCL8 which are associated to proinflammatory neutrophils and macrophage recruitment to the lungs.

**Figure 3.**
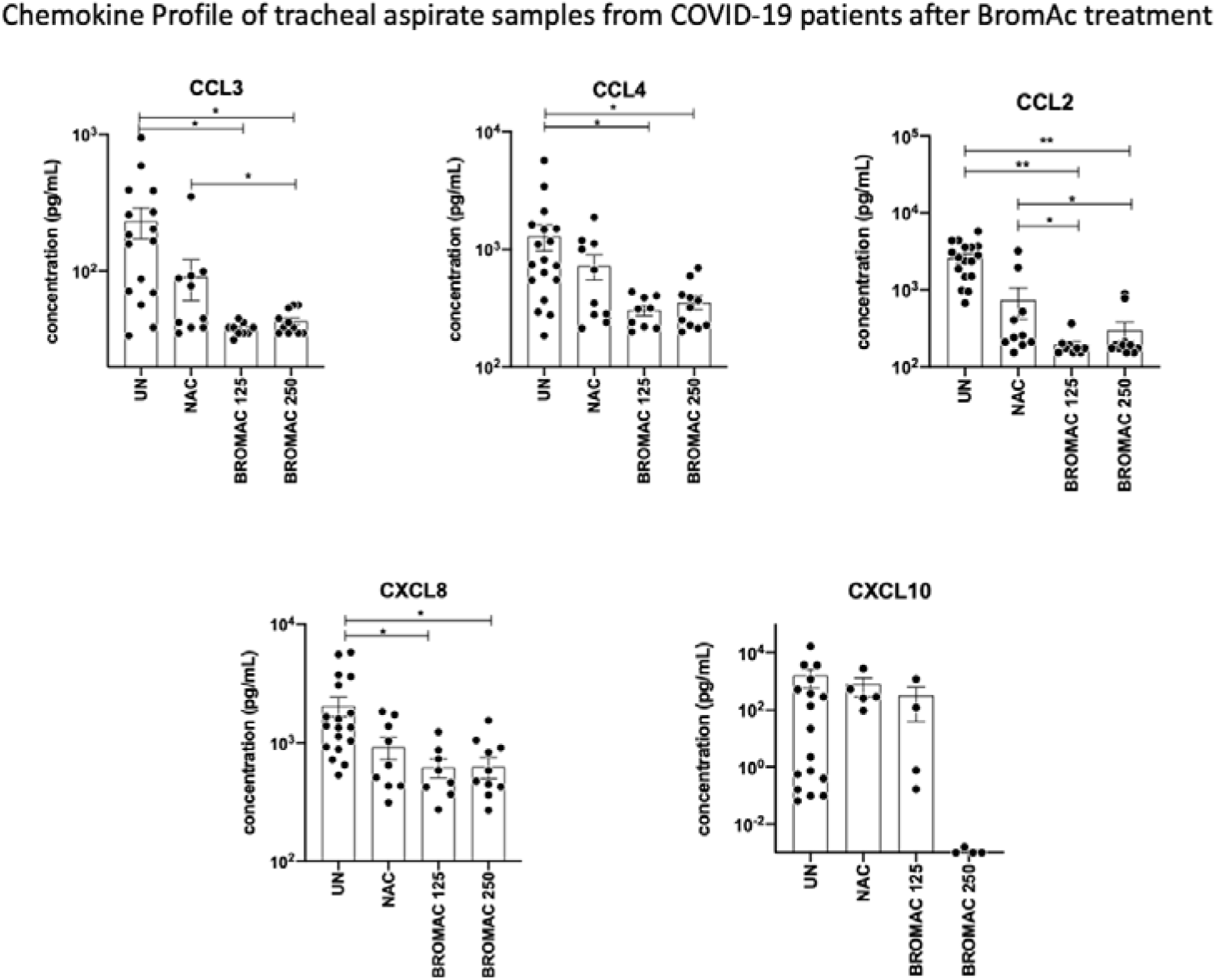
Effect of BromAc^®^ in Chemokines present in tracheal aspirate sample from COVID-19 patients. Chemokine results were measured by Luminex and are plotted as scatter graphs over bars expressing average and standard deviation. Sputum from 10 COVID-19 patients were examined. Results are expressed in pg/mL and statistical differences at p<0.05 were expressed as connecting lines and (*).

Furthermore, the cytokine storm was generally altered by BromAc^®^ treatment, showing lower levels of IL-6, a major inflammatory player on COVID-19 alongside IL-15, IFN-γ and IL-17A (Figure 4). Decreased levels of proinflammatory cytokines, IL-1β in association with IL-6 were observed after treatment with BromAc^®^, indicating possible modulation of inflammassome-associated pathway. IL-1β and TNF were also decreased in samples treated with BromAc^®^.

**Figure 4.**
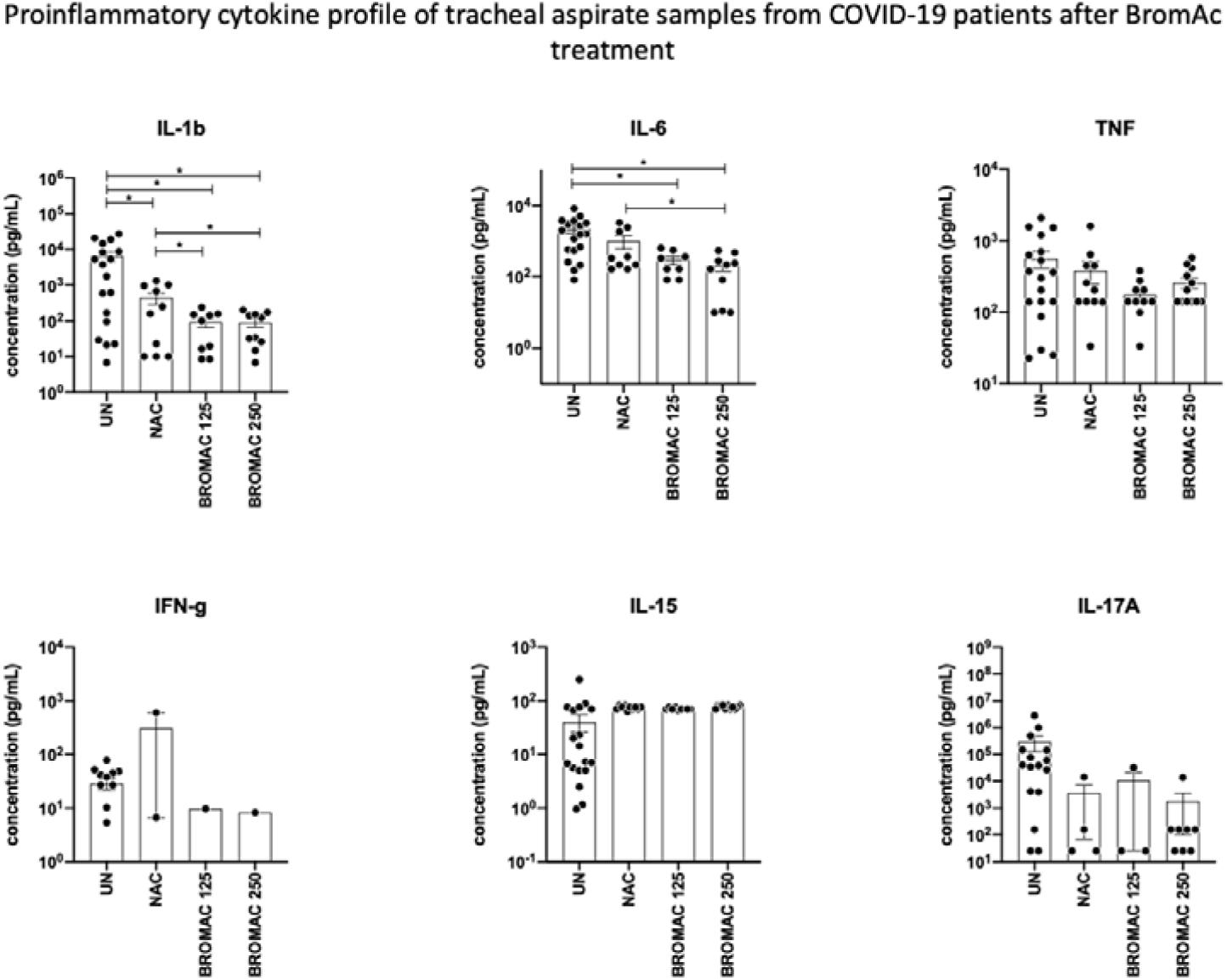
Effect of BromAc^®^ in pro-inflammatory cytokines present in tracheal aspirates from COVID-19 patients. Cytokine results were measured by Luminex and are plotted as scatter graphs over bars expressing average and standard deviation. Sputum from 10 COVID-19 patients was examined. Results are expressed in pg/mL and statistical differences at p<0.05 were expressed as connecting lines and (*).

The analysis of the ex vivo activity of BromAc^®^ on growth factors in the tracheal aspirate samples from COVID-19 patients indicated decrease levels of and vascular endothelial growth factor, VEGF-A. However, other growth factors such as VEGF-D, IL-2, PDGF and GM-CSF were increased after treatment with BromAc^®^ (Figure 5). Increased PDGF, a growth factor associated with platelet recruitment, should be better studied, but it could be associated to the degradation of platelets, which leads to the release of this growth factor. The same hypothesis could be drawn by other growth factors that could be membrane bound and detach from cells after BromAc^®^ treatment.

**Figure 5.**
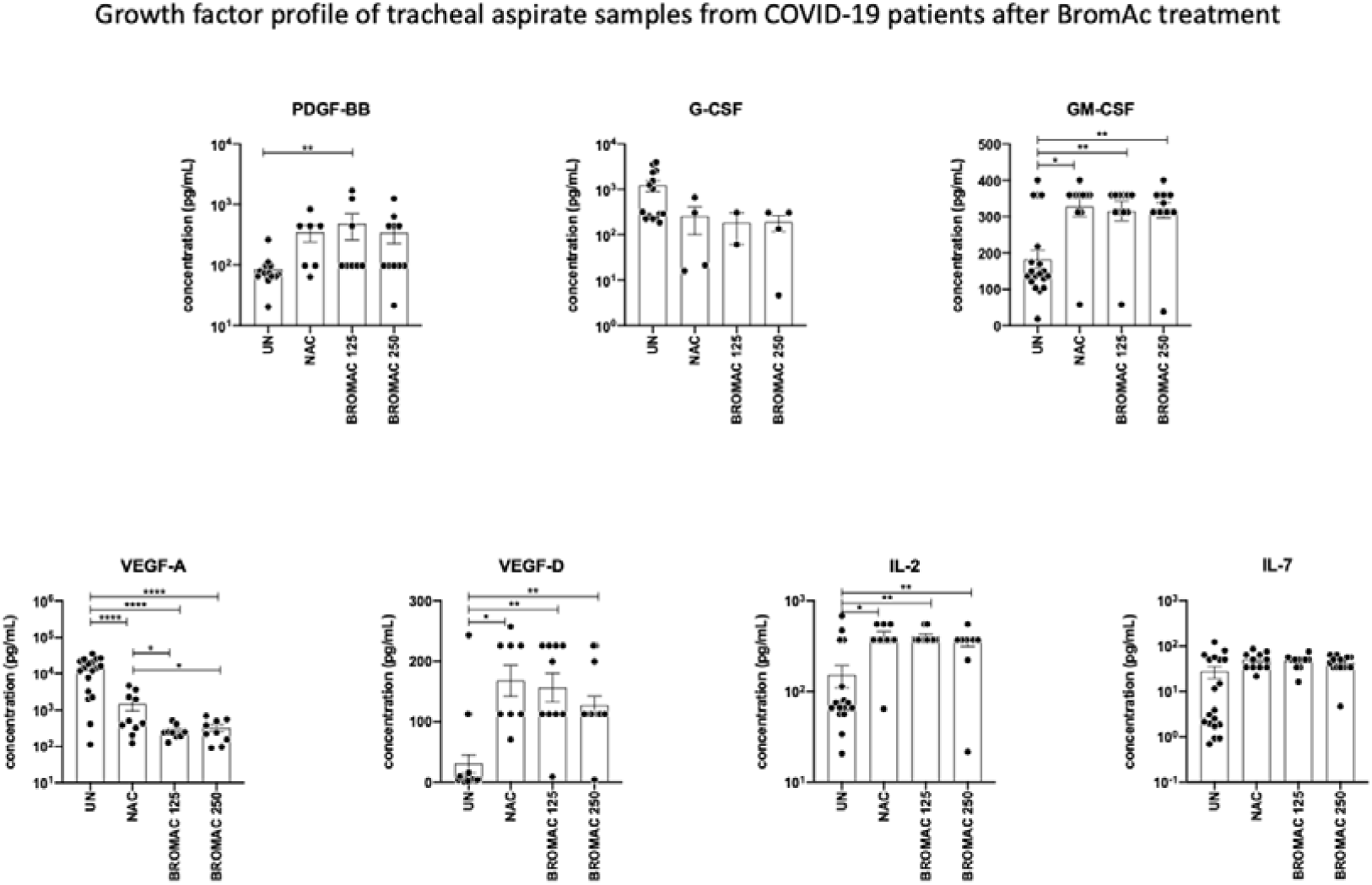
Effect of BromAc^®^ on growth factors present in tracheal aspirates from COVID-19 patients. The results for growth factors were measured by Luminex and are plotted as scatter graphs over bars expressing average and standard deviation. Sputum from 10 COVID-19 patients were assessed. Results are expressed in pg/mL and statistical differences at p<0.05 were expressed as connecting lines and (*).

Regarding the regulatory cytokines, the results show major abrogation of these molecules (Figure 6). IL-1RA levels were not significantly altered by BromAc^®^ treatment as compared to NAC. Conversely, decreased levels of IL-9 and IL-10 were observed when BromAc^®^ groups when compared to untreated tracheal aspirate samples. For IL-9, the same effect was observed in NAC-treated group, nevertheless, for IL-10, the significant decrease in the levels of this cytokine was solely observed in the BromAc^®^ groups. Excessive regulation may be associated or may lead to decreased responsiveness, anergy and diminished effector responses against the virus. Therefore, these results may indicate that BromAc^®^ could contribute to maintaining full effector responses, such as Type 1 immune responses against SARS-CoV-2 during the viral phase.

**Figure 6.**
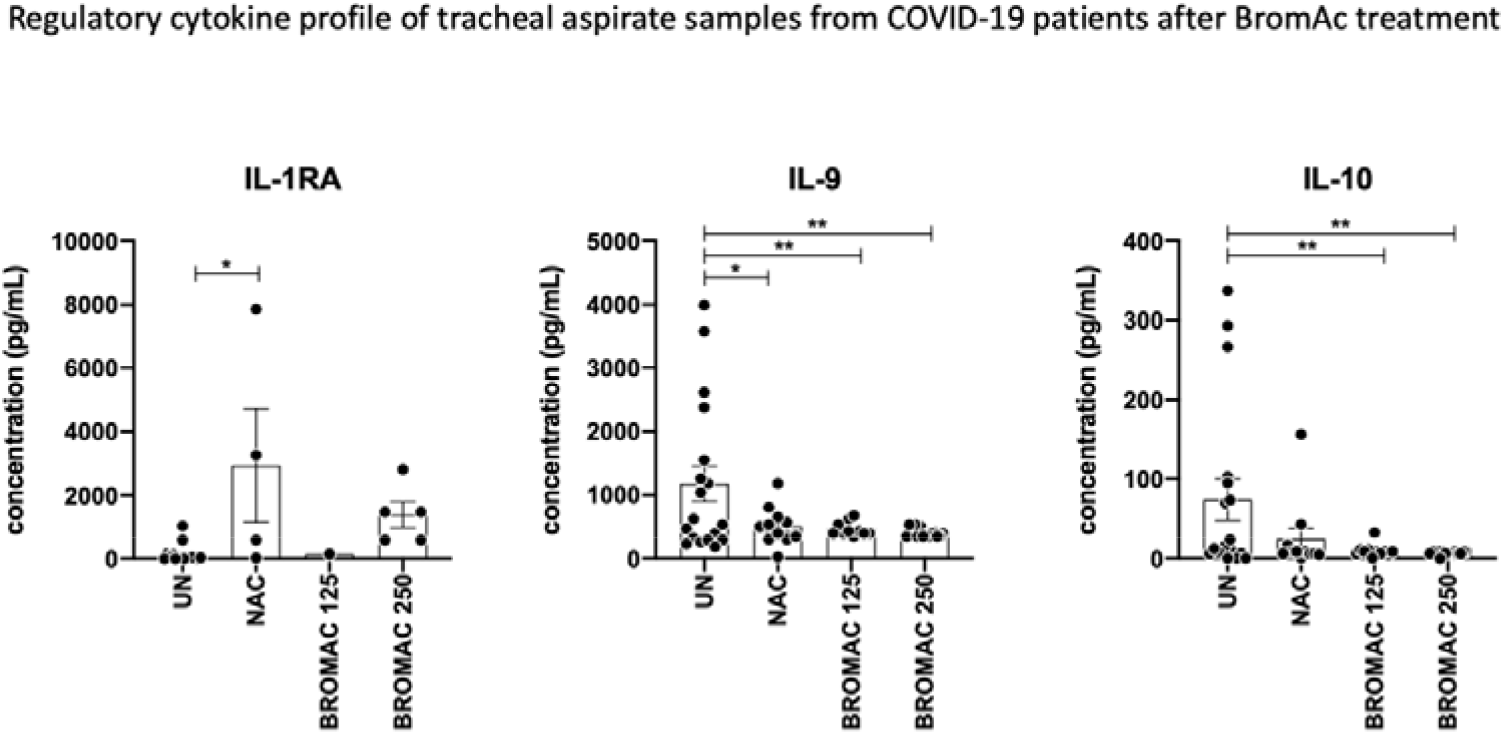
Effect of BromAc^®^ in regulatory cytokines present in tracheal aspirates from COVID-19 patients. Cytokine results were measured by Luminex and are plotted as scatter graphs over bars expressing average and standard deviation. Data from 10 COVID-19 patients were tested. Results are expressed in pg/mL and statistical differences at p<0.05 were expressed as connecting lines and (*).

## Discussion

There are many pathological changes in the lungs in COVID-19, and these evolve over time. Early disease is characterised by neutrophilic exudative capilliaritis with thrombosis. Late changes occurring on average from around day 10 include diffuse alveolar damage, intravascular thrombosis, infection, disseminated intravascular coagulopathy (DIC) and later intra-alveolar fibroblast proliferation [9].

Whilst COVID-19 is a multi-system disease, respiratory failure is a prominent cause of hospitalisation and death. Lung pathology in COVID-19 is complex and variable and time dependent, sputum plugging of airways is seen in a significant proportion of patients and adversely affects ventilation. The character of the sputum in COVID-19 is also highly viscous [10] and a higher proportion of critically ill COVID patients had grade 3 sticky sputum [11]. Bronchoscopy has revealed extensive mucus plugging or mucus in 64% [12] and 82% [13] and improvement in oxygenation after clearance. Mucus plugs were an adverse prognostic factor [13]. Endotracheal tube obstruction due to mucinous sputum is also a problem recognised far more frequently in COVID-19 patients than others requiring ventilation [14].

Sputum characteristics in patients with severe COVID-19 correlate with outcome. Wang et al. [15] described increasingly sticky sputum was associated with critical illness, being the viscosity, the major variable relating to sputum clearance. Highly viscous sputum in COVID-19 explains the high pressures required for ventilation and difficulty in suctioning COVID-19 sputum [16]. Therefore, effective expectorant drugs are strongly recommended during airway management [16].

Alveolar mucus affects the blood-gas barrier, inducing hypoxia, and provides direct evidence of the role of mucus in respiratory diseases and the mechanism we seek to disrupt with BromAc^®^. Therefore, for the first time, we evaluated the effect of BromAc^®^ in COVID-19 sputum as a strategy do reduce the sputum viscosity and enhance the airway management and the patient prognostic.

The treatment of COVID-19 sputum with BromAc^®^ enhanced the flow through (Figure 1A) and efficiently reduced the viscosity (Figure 2A-D) when compared to untreated samples or samples treated with NAC alone. All samples (untreated, NAC and BromAc^®^) showed a viscoelastic profile (Figure 2B), which is in according with the literature [7,17]. The cellularity analysis (Figures 1 C-D) showed an inverse correlation with the mucolytic effect of BromAc^®^. The viscoelastic fluid behaviour of mucus is directly proportional to the molecular weight and concentration of its components as well as their spatial configuration [18]. A recent contribution reports high solids and proteins in COVID-19 sputum similar to that seen in cystic fibrosis and identifies high levels of DNA and hyaluronan [19]. Our studies demonstrate that the use of BromAc^®^ provides a large reduction on viscosity of COVID-19 sputum at the first minute (Figure 2C), which we predict will be beneficial in improving pulmonary compliance on mechanical ventilation and improving sputum removal. This greater and rapid reduction in the viscosity of COVID-19 sputum can be explained by the synergistic effect of the two proteolytic drugs (NAC and bromelain) present in BromAc^®^, which enhanced the proteolytic effect.

A number of theoretical and experimental studies have demonstrated that the increase in viscosity and surface tension of airway surface liquid are likely responsible for ventilator induced lung injury (VILI). Two main physical mechanisms for VILI are lung tissue overdistention caused by surface tension-induced alterations in interalveolar micromechanics and atelecto-trauma the epithelial cells during repetitive airway reopening and closure [16].

There is a strong link between viral infection and mucus production via multiple signaling pathways including IL6, 1L10 and TNF whereby the cytokine storm causes sudden mucus hypersecretion [20]. Although viral infection can directly cause of excess sputum production by respiratory cells, the cytokine storm causes over production of mucus via both STAT MAPK, NKKB pathways in cells [20], and IFN-AhR signalling pathways in COVID [21]. Mucus production in respiratory diseases is normally related to inflammation and in COVID-19, it is established knowledge that mucin is induced by interferon [21] discovered in the bronchoalveolar lavage fluid (BALF) of COVID-19 patients and animals.

In severe cases, cytokine storm is due to the excess production of inflammatory cytokines including IL (interleukin)-1, IL-6, IL-12, IFN (interferon)-y and TNF (tumour necrosis factor)-∝ [22]. Increased serum levels of IL-7, IL-10, macrophage colony stimulating factor (CSF) (M-CSF), granulocytes CSF (G-CSF), granulocyte-macrophage (GM-CSF), 10KD interferon gamma induced protein (IP-10), monocyte chemoattractant protein (MCP-1), macrophage inflammatory protein (MIP-1OC) and tumour necrosis factor (TNF) are seen, especially in patients with severe disease. [23].

Cytokines are polypeptides and, therefore, a substrate for bromelain breakdown. However, there is added complexity in that bromelain can stimulate [24] or inhibit cytokine release from cells in different settings [25]. Bromelain reduced G-CSF, GMCSF, IFN-γ, MIP and TNF by inflamed tissue in IBD [26]. Bromelain can reduce neutrophil migration to inflammation [27] and whilst this may be related to reduced cytokines, it is also likely to be due to the removal of cell surface receptors on neutrophils, including CD44, CD62 and chemokine receptors [28] and has similar action in tumour cells [29].

High serum cytokine levels in COVID-19 are clearly related to severity of illness and risk of death. Cytokine storm is thought to be responsible for the increased bronchial mucus secretions in COVID-19 [20]. Perhaps of most relevance to COVID-19, Bromelain reduced airway inflammation in an ovalbumin induced inflammation mouse model [30] with a reduction in leucocytes and eosinophils in bronchoalveolar lavage, cellular lung infiltrate and serum interleukins IL-4, IL-12, IL-17 and IFN-∝.

Bromelain administered intraperitoneally has been shown to attenuate the development of allergen airway disease in a mouse model of ovalbumin induced allergic airway disease [31]. Bronchoalveolar lavage leukocytes and cytokines, lung histology and airway hyper-responsiveness were measured. Bromelain treatment resulted in reduced BAL leukocytes, eosinophils, CD4+ and CD8+ T lymphocytes, CD4+/CD8+ T cell ratio and IL-13. Similarly, Bromelain had an anti-inflammatory effect on nuclear factor kappa B (NFKB) and mitogen-activated protein kinases (MAPK) signal pathways in lipopolysaccharide stimulated RAW 264.7 macrophage cells in a dose dependent manner [32]. Due of its anti-inflammatory effects, Bromelain has been suggested as a treatment for COVID-19 [33]. Bromelain alone has been shown to have a direct antiviral effect on the SARS-CoV-2 virus, which causes COVID-19, likely due to its effect on the spike protein or the receptors utilised by the virus for host entry [34]. This research was performed in Vero E6 cells and required significantly higher doses than combination BromAc^®^. Bromelain has also been used in a viral pneumonia model with Semliki Forest virus [35,36].

Acetylcysteine is a widely used generic, administered routinely for paracetamol overdose [37]. It has other indications, including renal protection but is widely used in respiratory diseases as an orally administered mucolytic [38]. Acetylcysteine acts as a mucolytic due to its effect on disulphide bonds [39]. It has been applied in cystic fibrosis and bronchitis [38,40,41] however has been reported to produce mucorhoea from nebulising a 5-10% solution, which is hyperosmolar [37]. A 2% solution (or 20mg/ml) is recommended. In a clinical study of 10 patients undergoing thoracic surgery, Acetylcysteine was nebulised post-operatively. This was a blinded crossover study. Acetylcysteine was found to reduce sputum viscosity and increased the volume of sputum expectorated and oxygenation compared to control (saline) [42].

BromAc^®^ was developed to dissolve the highly mucinous tumour pseudomyxoma peritonei and its mechanism of action surrounds the synchronous breaking of peptide, glycoprotein and SS bonds [43]. It is already known to remove a range of MUC types from cancer cells [44], which is the basis for its potential in mucin-secreting respiratory diseases. As a new drug combination, it has now been applied in over 40 patients at much higher concentrations than used in this study, albeit intraperitoneally, and is entering Phase II trials in EU and USA for the rare mucinous tumour [45]. Animal safety experiments of airway administration have been completed in two species over three models with no evidence of toxicity at the concentrations examined.

Akhter et al [4] previously reported a synergistic inhibition of the infectivity of two strains of SARS-CoV-2 cultivated in Vero, BGM and CALU-3 cells, showing an antiviral effect of 4 log reduction by BromAc^®^. It remains to be understood whether BromAc^®^ might act as an antiviral or be a useful intervention for this action in later stage disease. However, the results from this study clearly indicate a robust mucolytic and anti-inflammatory effect of BromAc^®^ in tracheal aspirates from critically ill COVID-19 patients, supporting its potential as a therapeutic strategy for COVID-19. Clinical trials are planned to determine the safety and efficacy of BromAc^®^ in treating the disease that has been the biggest challenge of the 21st century, COVID-19.

## Data Availability

All data produced in the present study are available upon reasonable request to the authors

## Acknowledgments

We thank our hospital and university colleagues and administrators for their support of this project. We also thank the PDTIS FIOCRUZ Flow Cytometry core facility for technical support on performing Luminex assay and analysis. We are thankful to all funding agencies that supported our work.

## Author contributions

JGACR, SJV, DLM, LDC, CAQ designed and conceived the study. HPPF, NRROJ, GRA, BMM, APMM, MMS, RF, LJM, RSAC, ATSL, FBF, RPR, JCTD, EG, CAQ selected patient’s sample, prepared paperwork and clinical data. CPSMM, MFS, MMDCS, SRG, GRA, BMM, URS, ITSL, HFF, RPR performed diagnosis by RT-PCR and designed molecular assays. GACG, JGACR, MGS, DPAM, LLF performed rheologic analysis, JGACR, GMF, PMO, FAC, ALR performed immunological assays and Luminex. JGACR, GMF, PMO, FAC, ALR, GACG, MGS analyzed data and prepared graphical data. JGACR, SJV and DLM prepared figures and wrote the manuscript. All authors reviewed and edited the final version of the manuscript.

